# Cultural Adaptation and Psychometric Assessment of Acceptability, Appropriateness, and Feasibility Measures for Family Planning Interventions Among Midwives and Nurses in Ghana

**DOI:** 10.64898/2026.03.16.26348483

**Authors:** Ernest Maya, Chris Guure, Franklin N. Glozah, Sarita Sonalkar, Arden McAllister, Roseline Doe, James Kiarie, Mary Eluned Gaffield

## Abstract

**Background:** Implementation research relies on accurate measurement of early implementation outcomes such as acceptability, appropriateness, and feasibility. However, instruments developed in high income settings may demonstrate limited cross-cultural validity when applied in different health systems. In postpartum family planning, differences in counselling routines, workload, and service organisation mean that unadapted measures can overlook important aspects of provider experience. This study aimed to culturally adapt and psychometrically evaluate the Acceptability of Intervention Measure, the Intervention Appropriateness Measure, and the Feasibility of Intervention Measure when applied to postpartum family planning counselling and use of the World Health Organization Medical Eligibility Criteria mobile application among midwives and family planning nurses in Ghana.

**Methods:** A sequential exploratory mixed methods design was employed. Three focus group discussions with 18 midwives and family planning nurses were conducted to culturally adapt the original measures developed by Weiner and colleagues. Insights from the qualitative phase informed revisions to the survey instrument. The adapted tool was subsequently administered to 150 midwives who had used the Medical Eligibility Criteria mobile application during one-on-one postpartum family planning counselling. Psychometric evaluation included assessment of internal consistency reliability, exploratory and confirmatory factor analysis, predictive validity, and tests of convergent and discriminant validity.

**Results:** The adapted implementation outcome measures demonstrated strong internal consistency, with Cronbach alpha coefficients ranging from 0.82 to 0.93 across the three constructs for both the counselling intervention and the mobile application. Confirmatory factor analysis indicated acceptable model fit following minor modifications, supporting the unidimensional structure of the acceptability, appropriateness, and feasibility constructs. Predictive validity was demonstrated through significant associations between implementation outcome scores and providers’ preferences regarding counselling practices and use of digital tools. Although the expanded adapted scales showed high reliability, only the abridged versions consisting of four acceptability items and three items each for appropriateness and feasibility demonstrated full convergent and discriminant validity.

**Conclusion:** Culturally adapted versions of the Acceptability of Intervention Measure, Intervention Appropriateness Measure, and Feasibility of Intervention Measure can be reliably applied to evaluate postpartum family planning interventions among midwives and family planning nurses in Ghana. The findings suggest that the abridged versions of these measures retain stronger construct validity than expanded adapted versions, highlighting the importance of balancing contextual adaptation with preservation of the underlying measurement structure.

**Contributions to the literature:** This study advances the field of implementation science by demonstrating the process and value of culturally adapting and validating the acceptability, appropriateness and feasibility measures for use in a low- and middle-income country context. It provides empirical evidence supporting the reliability and construct validity of these measures in assessing the implementation of postpartum family planning interventions, including mobile health applications. The findings contribute to the broader goal of strengthening implementation measurement strategies in resource-constrained settings and offer a replicable model for adapting psychometric tools across diverse healthcare environments.

## Background

Mobile health approaches are increasingly being used to strengthen service delivery across maternal, newborn, and reproductive health settings, particularly in low- and middle-income countries where frontline providers often work under conditions of limited time, staff shortages, and uneven access to decision support tools (1,2). Digital tools such as mobile applications are intended to improve the consistency, quality, and timeliness of care, but their success depends not only on technical functionality but also on how they are experienced by health workers during routine practice. Within implementation science, acceptability, appropriateness, and feasibility are recognised as important early implementation outcomes because they help explain whether an intervention is perceived positively, judged to fit the practice setting, and considered practical to use in routine care (3). These outcomes are especially important in formative and pilot implementation work, where researchers seek to understand whether an intervention is likely to be adopted and sustained. Reliable and valid tools for measuring these outcomes enable meaningful evaluation and comparison of implementation strategies. To support such measurement, Weiner and colleagues developed three psychometric tools: the Acceptability of Intervention Measure (AIM), the Intervention Appropriateness Measure (IAM), and the Feasibility of Intervention Measure (FIM) (4). These scales are frequently used in formative and pilot studies; however, when applied in new cultural or clinical settings, scales often require adaptation to ensure conceptual equivalence and measurement accuracy. Sound psychometric evaluation is essential when implementation outcome measures are applied in new service settings because reliability alone does not establish whether items retain their intended meaning across contexts. Evidence on internal consistency, factor structure, and construct validity is needed to determine whether a measure is capturing the same underlying concepts in a different health system, provider group, or intervention context.

The reliability, validity, and pragmatic measures of these outcomes are important for both monitoring and evaluation of the implementation efforts and their effectiveness (5,6). This is consistent with the present implementation study examining the use of the World Health Organization Medical Eligibility Criteria for Contraceptive Use mobile application during one-on-one postpartum family planning counselling. Postpartum family planning has a favourable impact on maternal and child morbidity when it results in couples spacing their pregnancies at least two years apart (7,8). Providing women who had facility-based childbirth with contraceptives before they are discharged is crucial to reducing unintended pregnancies among postpartum women (9). Although modern contraceptives are safe and effective with existing guidelines for their delivery in the postpartum period (10), many postpartum women do not use modern contraceptives despite wanting to avoid pregnancy (11). This is due to concerns about side effects, safety, inconvenience, and misconceptions (12,13). While healthcare providers can address some of these concerns, they need appropriate education and tools. In accordance with this, the WHO developed the MEC App which was field tested in three health facilities in Ghana in addition to promoting one-on-one postpartum family planning counseling (14). (10) In Ghana, postpartum family planning counselling is delivered largely by midwives and family planning nurses, who play a central role in helping women make contraceptive decisions during a clinically and socially sensitive period. Their judgments about whether counselling approaches and digital decision support tools are acceptable, appropriate, and feasible are therefore central to successful implementation. Understanding these perceptions is especially important in busy public sector settings where workflow demands, counselling norms, and provider client interaction patterns may differ from those in the settings where these measures were originally developed.

These measures were selected because they are among the most widely used tools for assessing acceptability, appropriateness, and feasibility in early implementation work. However, measures developed in high income settings may not function in the same way in settings with different service delivery arrangements, communication styles, provider expectations, and organisational realities. Without cross cultural adaptation, item wording may appear clear on the surface while still failing to reflect how providers interpret concepts such as suitability, practicality, or approval in their everyday work. Cross cultural adaptation is therefore necessary to strengthen conceptual equivalence, semantic clarity, and contextual relevance before psychometric performance is tested in a new setting. This approach aligns with widely recommended procedures for cross cultural adaptation of measurement instruments, which emphasise contextual review, expert consultation, and psychometric evaluation to ensure conceptual equivalence across settings (15,16)

As these instruments were originally developed in a different setting, adaptation and validation were necessary before they could be confidently applied in Ghana(17–19). The objective of this study was to culturally adapt and psychometrically assess the Acceptability of Intervention Measure, the Intervention Appropriateness Measure, and the Feasibility of Intervention Measure for evaluating one-on-one postpartum family planning counselling and the WHO Medical Eligibility Criteria mobile application in Ghana. Specifically, we evaluated the internal consistency, factorial validity, and construct validity of the adapted measures when applied to postpartum family planning counselling and use of the MEC App in Ghana.

## Methods

### Study design and settings

A sequential exploratory mixed methods design was employed. In this design, qualitative data were first collected to explore how midwives and family planning nurses interpreted the constructs of acceptability, appropriateness, and feasibility in the local context. Findings from the qualitative phase informed the cultural adaptation of the measurement items, which were subsequently evaluated through quantitative psychometric testing. This approach allowed context specific provider perspectives to inform instrument adaptation before testing the reliability and validity of the adapted measures.

The study was conducted in six hospitals across the Greater Accra and Eastern regions, all of which provide dedicated family planning services in addition to general and maternity services. The qualitative and quantitative components were implemented in different groups of hospitals for methodological and operational reasons. The three hospitals selected for the qualitative phase (Achimota, Shai Osudoku, and Pentecost) were comparable in level of care, staffing patterns, and family planning service delivery to the three hospitals where the MEC App was field-tested (Maamobi General Hospital, Ga West Municipal Hospital, and Nsawam Government Hospital). Conducting the qualitative work in comparable but separate facilities minimised contamination between phases while allowing the adaptation of the Acceptability, Intervention Appropriateness, and Feasibility measures to be informed by providers with similar training, caseloads, and service delivery responsibilities. The quantitative phase was then conducted exclusively in the MEC App test sites to ensure that psychometric evaluation reflected the experiences of providers who had directly used both the one-on-one counselling strategy and the mobile application.

The one-on-one counselling intervention consisted of structured postpartum family planning counselling provided by midwives using standard national guidance. The counselling covered postpartum fertility, available methods, eligibility, and screening for medical conditions. The MEC App was a mobile tool that guides providers in determining whether a woman is eligible for a specific contraceptive method. Providers entered basic clinical information into the app, which returned method specific eligibility guidance based on the World Health Organization criteria. In this study, the measures assessed how providers perceived using the counselling approach and the app during routine work, which clarified the interpretation of acceptability, appropriateness, feasibility, and predictive validity outcomes.

### 2.2 Study population

The study participants were midwives and family planning nurses who are the primary providers of family planning and postnatal services in Ghana. All students and rotation nurses and midwives were excluded from this study.

Providers were the focus of the study because acceptability, appropriateness, and feasibility are conceptualized in implementation research as provider-level outcomes during early implementation. The aim of the study was to understand how the one-on-one counselling approach and the MEC App performed in routine work from the perspective of those who deliver the interventions. Including clients would have required different design and client specific measures, which were outside the scope of this validation study.

## Qualitative study

### Sampling and sampling procedure

To explore, understand and modify the constructs and all the items in the original AIM, IAM, and FIM tool developed by Weiner and colleagues (14), we conducted three Focus Group Discussions (FGD); one each at the Achimota, Shai Osudoku and Pentecost hospitals in the Greater Accra region. These facilities are at the same level as the three facilities for field testing of the MEC App (Maamobi General Hospital, Ga West Municipal Hospital and Nsawam Government Hospital) and are staffed by the same level of health care providers.

Participants were eligible for inclusion if they were midwives or nurses who provided family planning counselling or postpartum services at the selected hospitals. Each FGD involved six purposefully selected midwives/family planning nurses (total sample of 18). The number of focus groups and participants was considered sufficient to achieve thematic saturation within a relatively homogeneous professional group, where participants shared similar training, responsibilities, and service delivery environments. All focus group discussions were conducted in English, which is the professional working language used by midwives and nurses in clinical practice in Ghana. Trained research assistants conducted the FGDs.

### Qualitative Data Analysis

The transcribed data were coded by trained research assistants. Framework analysis provided the overarching structure for organising the data, beginning with familiarisation, development of an analytic matrix, and systematic charting of responses across the predetermined domains of acceptability, appropriateness, and feasibility. Within this structured framework, thematic analysis was applied to inductively identify recurring patterns, nuances, and context-specific information that extended beyond the original AIM, IAM, and FIM constructs. This integrated approach allowed us to retain the conceptual boundaries of the AIM, IAM, and FIM constructs while generating culturally grounded themes that informed the adaptation and addition of scale items.

Themes identified during analysis were assessed to determine whether they represented discrete, actionable aspects of acceptability, appropriateness, or feasibility that were not adequately captured by the original AIM, IAM, and FIM items. Only themes that reflected (a) a clearly expressed perception held by multiple participants, (b) direct relevance to providers’ experience of delivering one-on-one counselling or using the MEC App, and (c) conceptual alignment with one of the three constructs were translated into candidate scale items. Themes that were anecdotal, idiosyncratic, or unrelated to the core constructs were excluded. Draft items derived from these themes were then reviewed by a family planning fellow and three experienced family planning providers who served as clinical leads for the postpartum family planning package. This review process served as an expert assessment of item clarity, relevance, and redundancy, although a formal Content Validity Index was not calculated. Their review focused on clarity, relevance, and redundancy with existing items. Based on their feedback, several items were refined for wording, merged to avoid overlap, or removed if judged duplicative or insufficiently distinct. The final adapted item set formed the basis for the quantitative psychometric evaluation. Although formal cognitive interviewing procedures were not conducted, semantic clarity and conceptual interpretation of the items were explored during the focus group discussions, where participants discussed how they understood each construct and the wording of potential survey items.

The decision to create new items was guided by whether a theme was expressed repeatedly across discussions and reflected a clear and consistent understanding of acceptability, appropriateness, or feasibility among providers. Themes were included when they captured elements not addressed by the original items, such as clarity of content, comfort during use, and the need for simple and reliable tools, and were excluded if they did not relate directly to the three constructs or appeared only once. Each draft item was reviewed by the research team to confirm conceptual alignment and avoid redundancy. This process resulted in the final set of culturally adapted AIM, IAM, and FIM items, which were incorporated into the survey instrument used for quantitative psychometric assessment. For example, providers frequently described acceptability in terms of comfort and personal approval when delivering counselling. This theme informed the item “Doing one on one counselling is comfortable.” Similarly, participants emphasized the importance of simplicity and manageable steps when using the mobile application, which informed feasibility items such as “The intervention is easy to use” and “The intervention is doable.” Themes that appeared only once or did not clearly align with the constructs of acceptability, appropriateness, or feasibility were not translated into scale items.

## Quantitative study

### Sample size determination

The sample size calculation was based on the formula proposed by Cochran (20). The anticipated proportion of study participants who would support one-on-one counselling and the use of the MEC App was assumed to be 50% due to lack of knowledge on the acceptability or otherwise of the tool in our setting and *e*=5% was the margin of error associated with the point estimates. A total sample size of 385 providers was determined. The sampling units were the individual midwives and family planning nurses in the three participating hospitals (Maamobi General Hospital, Ga West Municipal Hospital and Nsawam Hospital). Records from the three hospitals indicated that the total number of midwives and family planning nurses was 201, which was less than the 385 calculated.

Therefore, the Finite Population Correction Formula (FPCF) was used to adjust the sample size. This yielded a representative minimum sample size of 131. The total sample size after adjusting for 15% non-response was 150.

#### The Adapted AIM, IAM, and FIM Survey Tool

##### One-on-one Counselling

For one-on-one postpartum family planning counselling, the adapted survey comprised of three subscales measuring acceptability (12 items), appropriateness (7 items), and feasibility (8 items). The item wording and corresponding factor loadings are presented in Table 3.

All items were administered as parallel statements referring specifically to the one-on-one counselling intervention. Responses were recorded using a five-category ordinal scale coded as completely disagree (0), disagree (1), neither agree nor disagree (2), agree (3), and completely agree (4), with higher scores indicating more favourable perceptions. Item level scores were used to generate construct scores for reliability, factor, and validity analyses.

##### MEC App

For the MEC App, the adapted survey comprised three subscales measuring acceptability (13 items), appropriateness (7 items), and feasibility (7 items). The item wording and corresponding factor loadings are presented in Table 4.

All items referred specifically to use of the mobile application during counselling. Responses were recorded using the same five category ordinal scale coded as completely disagree (0), disagree (1), neither agree nor disagree (2), agree (3), and completely agree (4), with higher scores indicating more favourable perceptions. Item level scores were used to generate construct scores for reliability, factor, and validity analyses.

#### 2.4.3 Data collection procedure

Data collection was carried out by trained research assistants who approached all midwives and family planning nurses who provided care for postpartum women in the selected hospitals. Providers were eligible if they had used the one-on-one counselling approach and the MEC App during routine work and were present at post during the data collection period. Midwives and family planning nurses had used the MEC App during routine postpartum counselling activities in the participating facilities prior to completing the survey, ensuring that responses reflected practical experience with the intervention. A total of 150 midwives and family planning nurses participated in the study.

Data were collected electronically using REDCap, which allowed real time data entry and verification. The use of REDCap reduced the time required for data entry and processing and allowed the team to check entries during the period of data collection. Issues relating to completeness or accuracy were identified during the interviews and corrected immediately.

### Quantitative Data analysis

The data were analysed using AMOS (SPSS 22) and the R programming language. Basic descriptive measures such as proportions, means, percentages, were obtained depending on the scale of measurement of the variable. A reliability test was conducted using Cronbach’s alpha to examine the internal consistency of the items and their relationship with the constructs. A bivariate Pearson correlations analysis was also performed to assess the predictive validity of the outcome measures. Further, Factor Analysis was conducted using Exploratory Factor Analysis (EFA) and a Simultaneous Confirmatory Factor Analysis (SCFA) using a Maximum Likelihood estimation method with structural equation modelling. Psychometric evaluation involved exploratory factor analysis to examine the underlying dimensionality of the adapted items, followed by confirmatory factor analysis using structural equation modelling to test the expected three factor structure representing acceptability, appropriateness, and feasibility (21) A number of models were developed to test the hypothesis that a relationship between observed variables and their underlying latent constructs exist. As a result, a number of goodness-of-fit indices such as the chi-square test, the goodness of fit index (GFI), the root mean square error of approximation (RMSEA), the normed fit index (NFI), the non-normed fit index (NNFI) or the Tucker-Lewis index (TLI), and the comparative fit index (CFI) were evaluated. Finally, convergent and discriminant validity for each of the outcome measures of one-on-one counselling and the MEC App were assessed using: (1) Average Variance Extracted (AVE) - a measure of the amount of variance that is captured by a construct in relation to the amount of variance due to measurement error; (2) Maximum Shared Squared Variance (MSV) - the extent a variable can be explained in another variable and (3) Composite Reliability (CR) - the degree that a latent is explained by its observed variables. Discriminant validity is achieved when AVE is greater than MSV or average shared squared variance (ASV). For convergent validity, AVE should be equal or greater than the minimum threshold of 0.50 and lower than CR (22) As the available provider population was limited, the same sample was used for both exploratory and confirmatory factor analysis. While this approach is commonly used in scale development studies with limited samples, it does not allow independent cross validation of the measurement model.

### Ethical Considerations

The study was approved by the Ethics Review Committee of the Ghana Health Service (Protocol ID GHS-ERC 001/05/21). All the study participants provided written informed consent after they had been informed that their participation in the study was voluntary, and they could stop participating at any time without any consequences. They were also assured that no personal/identifiable information would be included in the report. The study posed minimal risk to participants because the survey did not involve supervisory evaluation or questions likely to cause emotional distress.

## 3 RESULTS

### 3.1 Qualitative Findings from Focus Group Discussions

Three focus group discussions explored how midwives and family planning nurses understood the acceptability, appropriateness, and feasibility of one-on-one counselling and the MEC App. These outcomes informed the cultural adaptation of AIM, IAM, and FIM items. Themes reflected providers’ interpretations of the constructs and their experiences delivering postpartum family planning services.

During the discussions, participants clarified how certain terms commonly used in implementation research were interpreted within their clinical practice vocabulary. For example, the term “fitting,” which appears in the original Intervention Appropriateness Measure, was commonly interpreted by participants as meaning “suitable for routine midwifery practice.” Similarly, the concept of “feasibility” was often described by participants in terms of whether an intervention was “doable” or “possible to implement during normal clinical work.” These interpretations informed minor wording adjustments to ensure that the adapted items reflected terminology familiar to providers.

#### 3.1.1 Acceptability

Participants described an acceptable counselling intervention as one that is collectively endorsed by staff, aligns with standard midwifery procedures, and is clearly explainable to clients. As one provider stated:

> “After the client has gotten whatever explanation, then the acceptance comes in.” *(Participant 6, Achimota)*

Training and availability of necessary tools were also central to acceptability:

> “I will accept an intervention if I have been trained well… and the tools needed are available.” *(Participant 2, Achimota)*

Providers linked acceptability to interventions that consistently produce positive outcomes:

> “After doing my intervention, I got what I needed, so it is acceptable.” *(Participant 6, Achimota)*

Practicality, comfort, and ease of use further shaped acceptability judgments:

> “It should be something that is comfortable for me.” *(Participant 5, Shai Osudoku)*

For the MEC App, acceptability depended on simplicity and accessibility:

> “An acceptable application should be one that a lot of people can use easily.” *(Participant 4, Shai Osudoku)*

> “Once you download it, no need for network… you can use it anywhere.” *(Participant 2, Shai Osudoku)*

#### 3.1.2 Appropriateness

Appropriateness reflected whether the intervention had the right content, served the intended purpose, and supported informed decision-making. Providers highlighted the need for clear, correct, and structured content in counselling:

> “It has the right content… it serves the right purpose.” *(Participant 3, Achimota)*

Counselling was considered appropriate when systematic and not overly time-consuming:

> “The intervention should be systematic… and it shouldn’t take too much time.” *(Participant 1, Shai Osudoku)*

Regarding the MEC App, providers emphasised its structured, stepwise process:

> “If you don’t complete this stage, you can’t move to the next… it streamlines you.” *(Participant 2, Pentecost)*

They also viewed the App as appropriate when it reinforced correct information and supported follow-up:

> “It helps with the type of information we are supposed to give.” *(Participant 3, Pentecost)*

> “It gives reminders for clients who need follow up.” *(Participant 4, Pentecost)*

#### 3.1.3 Feasibility

Feasibility was associated with practicability, accessibility, and comfort for both providers and clients.

> “It means something that can be done; you will be able to do it.” *(Participant 1, Achimota)*

> “Feasibility… makes me comfortable, and the client is also comfortable.” *(Participant 2, Achimota)*

Participants emphasised that feasible counselling is implementable across settings:

> “It can be done anywhere… it doesn’t come with any difficulty.” *(Participant 5, Shai Osudoku)*

For the MEC App, feasibility depended on reliable access, offline capability, and having manageable steps:

> “When you open the App and they tell you there’s network problem, you will find it difficult using it.” *(Participant 1, Shai Osudoku)*

> “When it can be used off data, it will make it feasible.” *(Participant 6 Pentecost)*

> “If the steps involved are very simple… but if there are too many steps, it won’t be feasible.” *(Participant 2, Shai Osudoku)*

### 3.2 Quantitative Findings

#### Socio-demographic Characteristics of Survey Participants

A total of 150 participants were included in the quantitative study. Majority of participants were between 32-37 years old (36.0%), followed by 26-31 years old (31.3%). Most participants had 1-5 years of experience in practice (41.3%), followed by 6-11 years (32.7%). The sample was predominantly married (73.3%) and identified as Christian, with the largest group belonging to the Church of Pentecost (50.0%). Almost all participants (99.3%) had attained a tertiary level of education. Table 1 shows the socio-demographic characteristics of survey participants.

**Table 1:** Socio-demographic characteristics of participating midwives and family planning nurses (N = 150)

| Socio-demographic variables | N | % |
| --- | --- | --- |
| **Age** |  |  |
| 26 ≤ age < 32 | 47 | 31.3% |
| 32 ≤ age < 38 | 54 | 36% |
| 38 ≤ age < 44 | 27 | 18% |
| 44 ≤ age < 50 | 7 | 4.7% |
| 50 ≤ age < 56 | 6 | 4% |
| 56 ≤ age ≤ 60 | 9 | 6% |
| **Number of years in practice** |  |  |
| 1-5 | 62 | 41.3% |
| 6-11 | 49 | 32.7% |
| 12-17 | 22 | 14.7% |
| 18-23 | 10 | 6.7% |
| 24-29 | 2 | 1.3% |
| 30-35 | 5 | 3.3% |
| **Marital status** |  |  |
| Single | 34 | 22.7% |
| Married | 110 | 73.3% |
| Divorced | 1 | 0.7% |
| Widowed | 4 | 2.7% |
| Separated | 1 | 0.7% |
| **Religion** |  |  |
| Catholic | 15 | 10% |
| Methodist | 12 | 8% |
| Jehovah Witness | 3 | 2% |
| Presbyterian | 23 | 15.3% |
| Church of Pentecost | 75 | 50% |
| Traditional | 1 | 0.7% |
| Islamic | 19 | 12.7% |
| *Other | 2 | 1.3% |
| **Highest level of education** |  |  |
| Tertiary | 149 | 99.3% |
| Tertiary-masters | 1 | 0.7% |

#### 3.2.1 Internal Consistency Reliability

The adapted measures demonstrated strong internal consistency for both the one-on-one counselling intervention and the MEC App. For one-on-one counselling, Cronbach’s α coefficients were 0.90 for acceptability, 0.82 for appropriateness, and 0.87 for feasibility. For the MEC App, Cronbach’s α values were 0.93, 0.86, and 0.90 respectively. Table 2 presents the reliability coefficients for both the adapted and original AIM, IAM, and FIM items. These values are comparable to, and in some cases higher than, the original AIM (α = 0.85), IAM (α = 0.91), and FIM (α = 0.89) reported by Weiner et al. (1). Notably, the appropriateness scale for one-on-one counselling closely aligned with the original IAM, while acceptability and feasibility showed modest increases, suggesting that the culturally adapted items resonated strongly with providers’ experiences.

**Table 2:** Cronbach alpha values for the adapted AIM, IAM, and FIM items and the original four item versions.

| Intervention and Measures | Current study |  | Original measures |  |
| --- | --- | --- | --- | --- |
| | Items (N) | $\alpha$ | Items (N) | $\alpha$ |
| **One-on-one counselling intervention** |  |  |  |  |
| Acceptability | 12 | 0.90 | 4 | 0.81 |
| Appropriateness | 7 | 0.82 | 4 | 0.83 |
| Feasibility | 8 | 0.87 | 4 | 0.82 |
| **MEC App** |  |  |  |  |
| Acceptability | 13 | 0.93 | 4 | 0.81 |
| Appropriateness | 7 | 0.86 | 4 | 0.84 |
| Feasibility | 7 | 0.90 | 4 | 0.86 |

#### 3.2.2 Factorial validity

To assess the measurement models of both the one-on-one counselling and MEC App, a Simultaneous Confirmatory Factor Analysis (SCFA), using a Maximum Likelihood estimation method with structural equation modelling, was performed, using their correlation matrix. For the one-on-one counselling measurement model (Figure 1), goodness-of-fit indices of the SCFA model did not show a good data fit, with χ^2^/df = 2.43, CFI = 0.77, TLI = 0.75, RMSEA = 0.10 - none of the fit indices matched the cut-off criterion. However, after correlating the error terms of variables i1 and i6 as well as variables i8 and i9, based on modification indices, the data had a good fit with the measurement model: χ^2^/df = 1.62, CFI = 0.96, TLI = 0.95, RMSEA = 0.06. The modified measurement model (Figure 2) had high and acceptable factor loadings (Table 3). Although the initial CFA model based on the original four-item AIM, IAM, and FIM structure was tested, the model-fit indices indicated poor fit to the one-on-one counselling data (χ²/df > 3, CFI < 0.80, TLI < 0.80, RMSEA > 0.10). These values were markedly below recommended thresholds, confirming that the original measurement structure did not adequately represent providers’ responses. Even after applying modification indices, the original four-item model did not reach acceptable fit, whereas the adapted multi-item model demonstrated substantially improved fit (χ²/df = 1.62, CFI = 0.96, TLI = 0.95, RMSEA = 0.06), as shown in Table 3.

**Figure 1.**
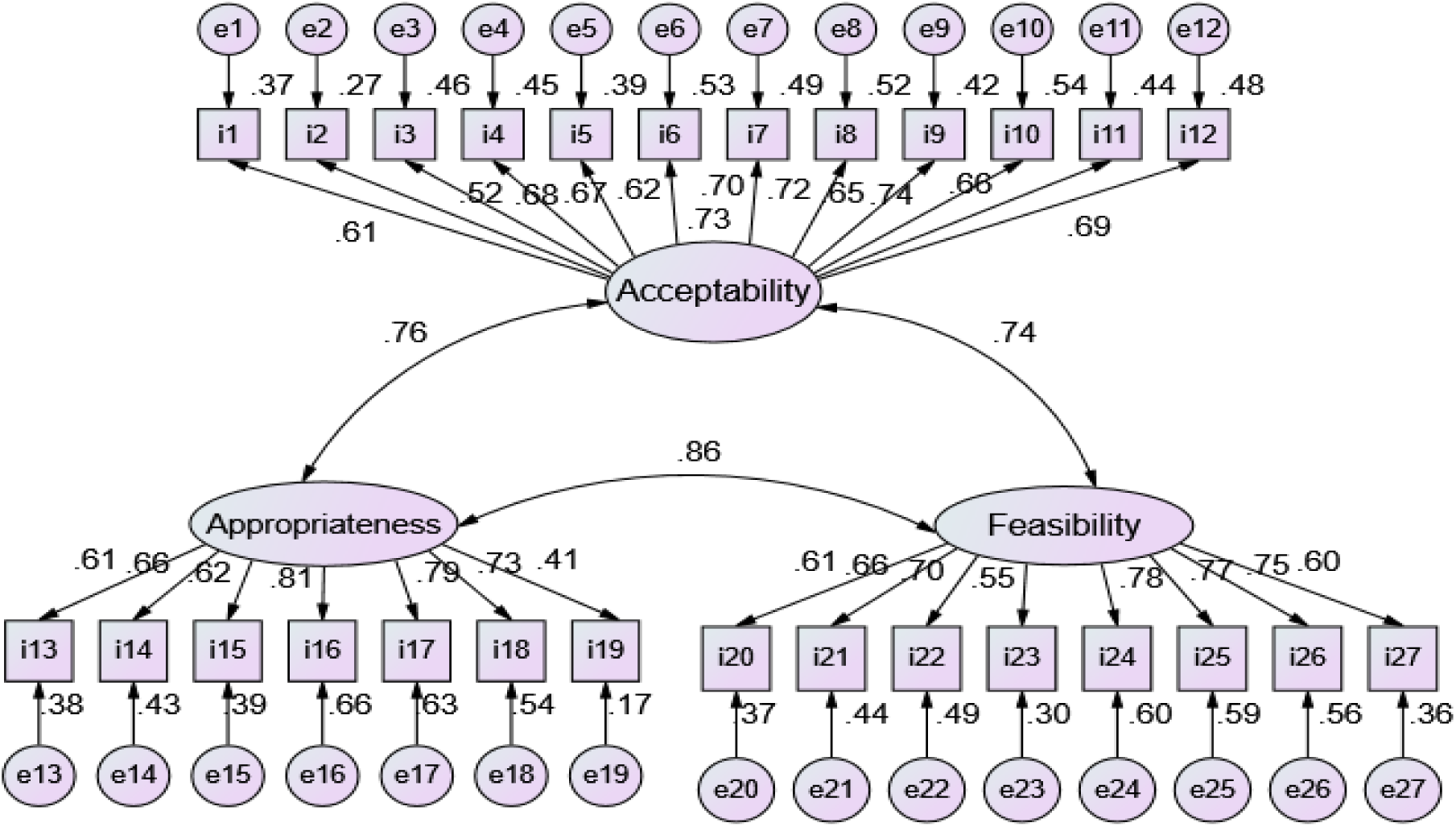
One-on-one counselling initial measurement model linking observed items to three latent variables

**Figure 2.**
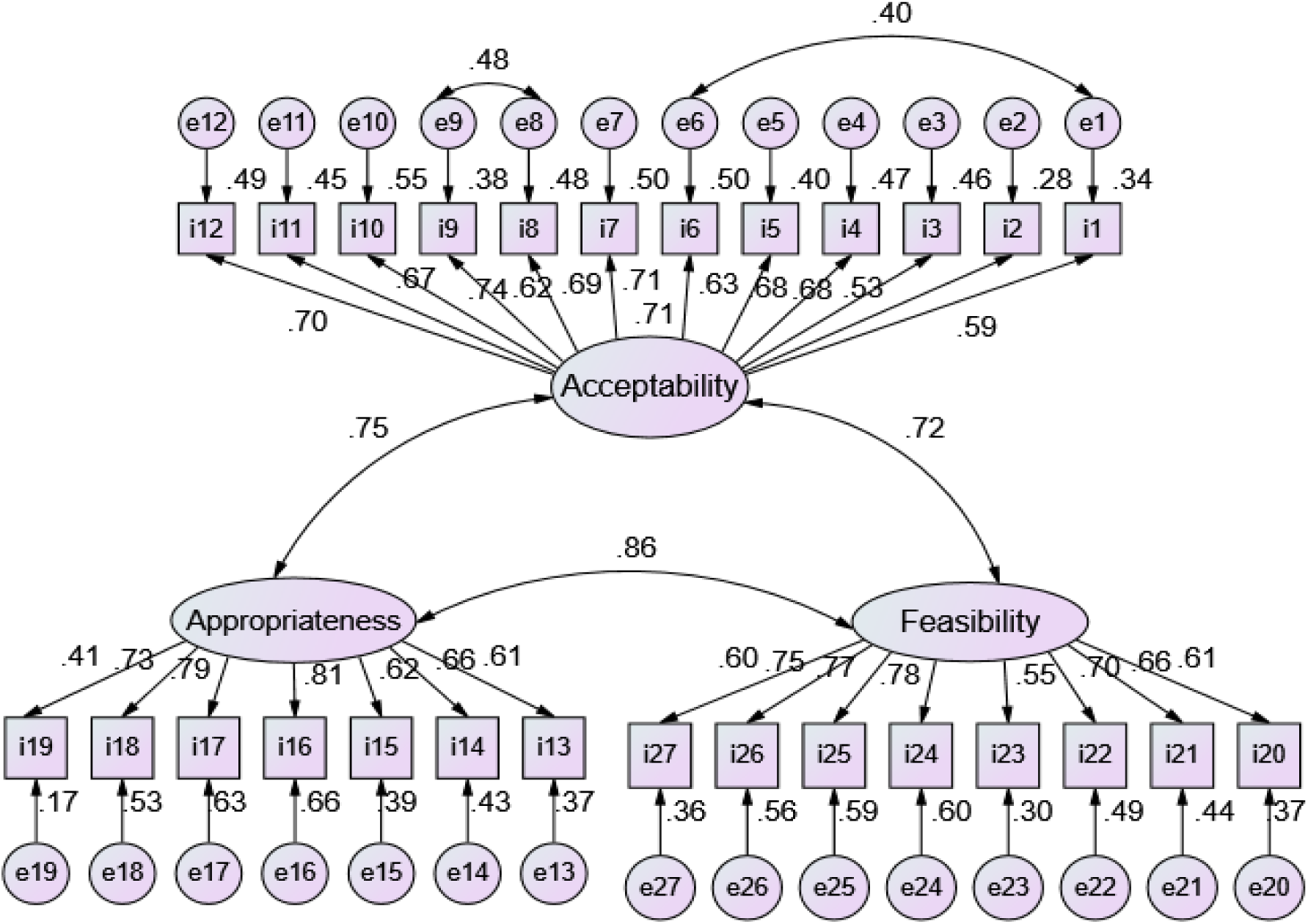
Modified one-on-one counselling measurement model linking observed items to three latent variables

**Table 3:** Standardised total effects (factor loadings) of the modified one-on-one counselling of the AIM, IAM and FIM.

| Item Number | One-on-one Counselling | Standardized Total Effects (Factor Loadings) |
| --- | --- | --- |
|  | **Acceptability** |  |
| i1 | **Doing one-on-one counselling meets my approval** | 0.70 |
| i2 | Doing one-on-one counselling allows for privacy | 0.67 |
| i3 | Doing one-on-one counselling is convenient | 0.74 |
| i4 | Doing one-on-one counselling is easy to understand | 0.62 |
| i5 | Doing one-on-one counselling meets standard of care | 0.69 |
| i6 | **Doing one-on-one counselling is appealing to me** | 0.71 |
| i7 | Doing one-on-one counselling is comfortable | 0.71 |
| i8 | Doing one-on-one counselling is attractive | 0.63 |
| i9 | Doing one-on-one counselling is negotiable | 0.69 |
| i10 | **I like doing one-on-one counselling** | 0.68 |
| i11 | **I welcome doing one-on-one counselling** | 0.53 |
| i12 | One-on-one counselling is workable | 0.59 |
|  | **Appropriateness** |  |
| i13 | **Doing one-on-one counselling is fitting** | 0.61 |
| i14 | One-on-one counselling is easy to explain | 0.66 |
| i15 | One-on-one counselling promotes specificity | 0.62 |
| i16 | **Doing one-on-one counselling is suitable** | 0.81 |
| i17 | **Doing one-on-one counselling is applicable** | 0.79 |
| i18 | **Doing one-on-one counselling is a good match.** | 0.73 |
| i19 | Doing one-on-one counselling takes a relatively short time | 0.41 |
|  | **Feasibility** |  |
| i20 | **Doing one-on-one counselling is implementable** | **0.61** |
| i21 | **Doing one-on-one counselling is possible** | **0.67** |
| i22 | **Doing one-on-one counselling is doable** | **0.70** |
| i23 | **Doing one-on-one counselling is easy** | **0.55** |
| i24 | Doing one-on-one counselling is practicable | 0.78 |
| i25 | Doing one-on-one counselling is applicable | 0.77 |
| i26 | Doing one-on-one counselling is understandable | 0.75 |
| i27 | Doing one-on-one counselling is convenient | 0.60 |

*Note: Items of the original outcome measures are in bold texts.*

**Table 4:** Standardised total effects (factor loadings) of the modified AIM, FIM, IAM measures used to evaluate the MEC App.

| Item Number | MEC App | Standardized Total Effects (Factor Loadings) |
| --- | --- | --- |
|  | **Acceptability** |  |
| **MA1** | **The MEC app meets my approval** | **0.79** |
| MA2 | The MEC app allows for privacy | 0.75 |
| MA3 | The MEC app is convenient | 0.76 |
| MA4 | The MEC app is easy to understand | 0.65 |
| MA5 | The MEC app is easy to use | 0.77 |
| MA6 | The MEC app meets standard of care | 0.73 |
| **MA7** | **The MEC app is appealing to me** | **0.70** |
| MA8 | It is comfortable to use the MEC app | 0.79 |
| MA9 | The MEC app is attractive | 0.71 |
| MA10 | The MEC app allows for negotiation | 0.71 |
| **MA11** | **I like the MEC app** | **0.75** |
| **MA12** | **I welcome the MEC app** | **0.67** |
| MA13 | The MEC app is workable | 0.64 |
|  | **Appropriateness** |  |
| **MA14** | **The MEC app is fitting** | **0.65** |
| MA15 | The MEC app is easy to explain | 0.75 |
| MA16 | The MEC app allows for specificity | 0.69 |
| **MA17** | **The MEC app is suitable** | **0.81** |
| **MA18** | **The MEC app is applicable** | **0.67** |
| **MA19** | **The MEC app is a good match** | **0.74** |
| MA20 | Doing one-on-one counselling with the MEC app takes a relatively short time | 0.63 |
| **Feasibility** |  |  |
| **MA21** | **The MEC app is implementable** | **0.79** |
| **MA22** | **The MEC app is possible** | **0.86** |
| **MA23** | **The MEC app is doable** | **0.73** |
| **MA24** | **The MEC app is easy to use** | **0.69** |
| MA25 | The MEC app is practical | 0.78 |
| MA26 | The MEC app is applicable | 0.73 |
| MA27 | The MEC app is understandable | 0.66 |

*Note: Items of the original outcome measures are in bold texts.*

Similarly, the MEC App measurement model (Figure 3), the goodness-of-fit indices of the SCFA model did not initially show a good fit with the data: χ^2^/df = 2.30, CFI = 0.85, TLI = 0.83, RMSEA = 0.09; none of the fit indices matched the cut-off criterion. Modification indices suggested correlating the following error terms: e4 <-> e5, e17 <-> e8 and e25 <-> e26. Indeed, this resulted in improved and acceptable data to model fit: χ^2^/df = 2.04, CFI = 0.88, TLI = 0.87, RMSEA = 0.08. Figure 4 shows the modified MEC App measurement model, which had high and acceptable factor loadings shown in Table 4. Similarly, the original four-item measurement model for the MEC App produced inadequate model-fit indices at baseline (χ²/df > 3, CFI < 0.85, TLI < 0.85, RMSEA > 0.10), indicating that it did not capture the underlying structure of midwives’ responses. Applying modification indices did not yield acceptable fit for the original model. In contrast, the adapted model incorporating culturally generated items showed improved and acceptable fit (χ²/df = 2.04, CFI = 0.88, TLI = 0.87, RMSEA = 0.08), with corresponding factor loadings presented in Table 4.

**Figure 3.**
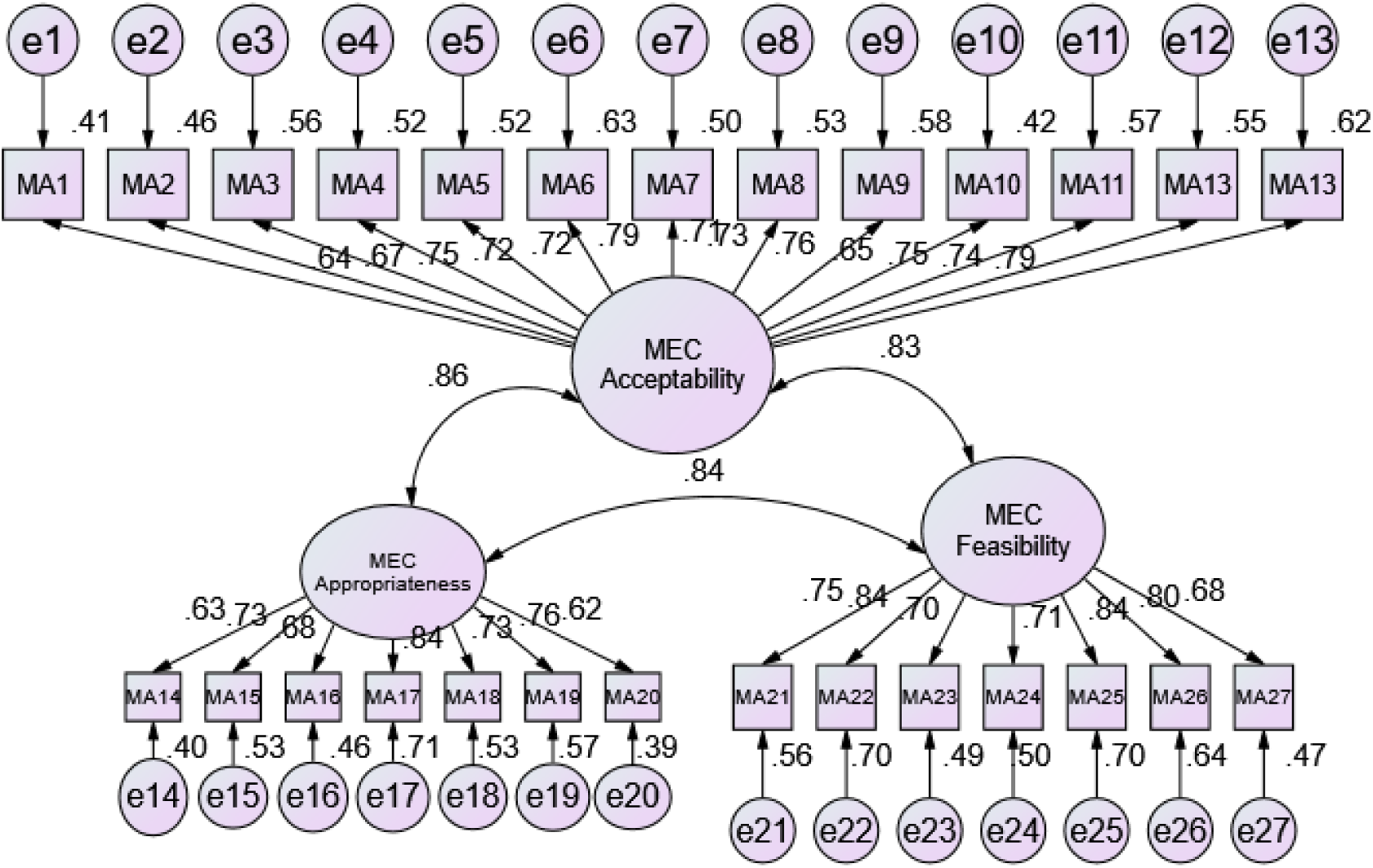
MEC App initial measurement model linking observed items to three latent variables

**Figure 4.**
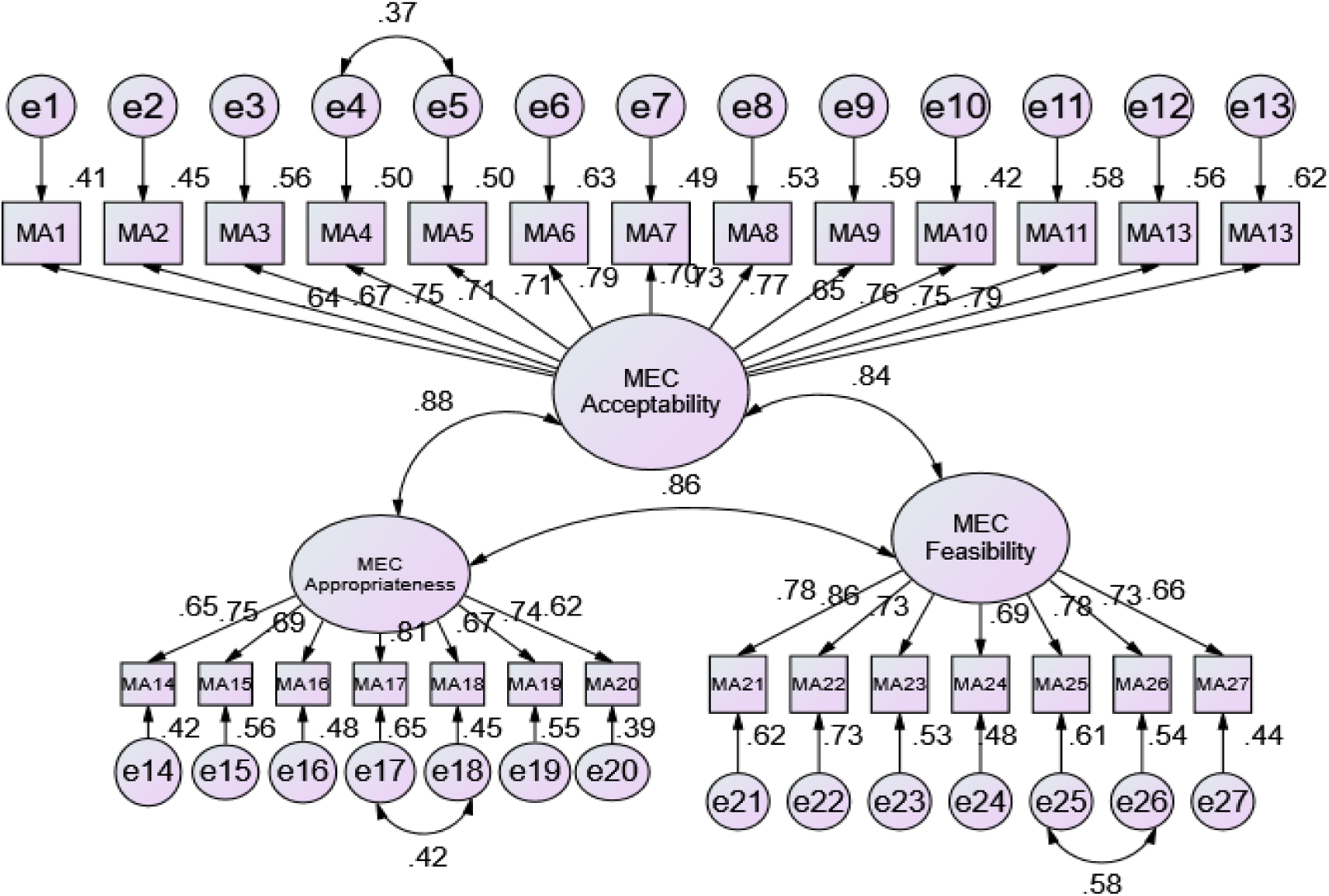
Modified MEC App measurement model linking observed items to three latent variables.

#### 3.2.3 Predictive validity

To test the predictive validity of both the one-on-one counselling intervention and MEC App regarding their acceptability, appropriateness, and feasibility, a bivariate Pearson’s correlations were performed (2-tailed), between factor scores of the AIM, IAM, and FIM measures and two predictive validity questions: “I prefer to work in a facility where one-on-one counselling is done,” and “Using computer applications in counselling would slow down the work.” It was expected that the factor scores of one-on-one counselling would be positively associated with the predictive validity question (7) while the factor scores of the MEC App will be negatively associated with predictive validity question (8) (Table 5).

**Table 5:** Correlation matrix of implementation outcome variables and predictive validity questions.

|  | 1 | 2 | 3 | 4 | 5 | 6 | 7 | 8 |
| --- | --- | --- | --- | --- | --- | --- | --- | --- |
| 1. Acceptability of one-on-one counselling | 1 |  |  |  |  |  |  |  |
| 2. Appropriateness of one-on-one counselling | .662** | 1 |  |  |  |  |  |  |
| 3. Feasibility of one-on-one counselling | .704** | .727** | 1 |  |  |  |  |  |
| 4. Acceptability of MEC App | .632** | .624** | .619** | 1 |  |  |  |  |
| 5. Appropriateness of MEC App | .604** | .671** | .618** | .790** | 1 |  |  |  |
| 6. Feasibility of MEC App | .657** | .557** | .626** | .768** | .759** | 1 |  |  |
| 7. I prefer to work in a facility where one-on-one counselling is done | **.496\*\*** | **.364\*\*** | **.404\*\*** | .474** | .513** | .498** | 1 |  |
| 8. Using computer applications in counselling would slow down the work | -.144 | -.109 | -.096 | **-.224\*\*** | **-.182\*** | **-.219\*\*** | -.075 | 1 |

\* $p < 0.05$ level (2-tailed), \*\* $p < 0.01$ level (2-tailed).

These questions were selected because they reflect practical judgments regarding the usefulness and efficiency of the interventions in routine clinical work.. A preference to work in a facility where one to one counselling is done reflects the expectation that the approach improves daily practice, while concern that computer applications slow down work reflects how providers judge the convenience and efficiency of the MEC App. These practical judgments are appropriate indicators for testing whether higher scores on acceptability, appropriateness, and feasibility are associated with favourable reactions to the interventions.

There was a strong positive correlation between the acceptability, appropriateness, and feasibility of one-on-one counselling and the predictive validity question, showing that providers who rated the counselling approach more positively were more likely to prefer working in a facility where it is offered. For the MEC App, higher acceptability, appropriateness, and feasibility scores were associated with lower agreement that computer applications slow down work. Together, these results show that the adapted scales performed well in predicting practical judgments related to each intervention.

#### 3.2.4 Convergent and discriminant validity

Convergent validity was evaluated using average variance extracted (AVE ≥ 0.50), composite reliability (CR ≥ 0.70), and standardised factor loadings (≥ 0.50). As presented in Tables 6 and 8, all adapted constructs for one-on-one counselling and the MEC App met these criteria. Most loadings were above 0.70, and all AVE values exceeded 0.50, indicating that items within each construct adequately captured a shared underlying concept. CR values were acceptable to high across all three constructs, providing further support for convergent validity.

**Table 6.** Convergent and discriminant validity of one-on-one counselling.

|  | **CR** | **AVE** | **MSV** | **1** | **2** | **3** |
| --- | --- | --- | --- | --- | --- | --- |
| 1. Acceptability | 0.81 | 0.52 | 0.49 | **0.72** |  |  |
| 2. Appropriateness | 0.84 | 0.64 | 0.44 | 0.58 | **0.80** |  |
| 3. Feasibility | 0.79 | 0.55 | 0.49 | 0.70 | 0.66 | **0.74** |

Notes: CR = Composite Reliability; AVE = Average Variance Extracted; MSV = Maximum Shared squared variance.

Discriminant validity was assessed by comparing the square root of each construct’s AVE with its inter-construct correlations. For both interventions, the square roots of the AVE were greater than the corresponding correlations between acceptability, appropriateness, and feasibility. This demonstrates that each construct was empirically distinct and measured a different dimension of providers’ perceptions. In both cases, the adapted measures showed strong convergent and discriminant validity in this context. Tables 7 and 9 shows results of the final number of items that resulted in an acceptable convergent and discriminant validity.

**Table 7.** Items of the original tool that produced acceptable convergent and discriminant validity of one-on-one counselling.

| Item | **One-on-one Counselling** | **Factor Loadings** |
| --- | --- | --- |
|  | **Acceptability** |  |
| 1 | Doing one-on-one counselling meets my approval | 0.68 |
| 2 | Doing one-on-one counselling is appealing to me | 0.78 |
| 3 | I like doing one-on-one counselling | 0.76 |
| 4 | I welcome doing one-on-one counselling | 0.67 |
|  | **Appropriateness** |  |
| 1 | Doing one-on-one counselling is suitable | 0.82 |
| 2 | Doing one-on-one counselling is applicable | 0.84 |
| 3 | Doing one-on-one counselling is a good match. | 0.73 |
|  | **Feasibility** |  |
| 1 | Doing one-on-one counselling is implementable | 0.66 |
| 2 | Doing one-on-one counselling is possible | 0.81 |
| 3 | Doing one-on-one counselling is doable | 0.75 |

**Table 8.** Convergent and discriminant validity of the MEC App.

|  | CR | AVE | MSV | 1 | 2 | 3 |
| --- | --- | --- | --- | --- | --- | --- |
| 1. Acceptability | 0.80 | 0.57 | 0.55 | **0.76** |  |  |
| 2. Appropriateness | 0.86 | 0.68 | 0.65 | 0.86 | **0.82** |  |
| 3. Feasibility | 0.87 | 0.70 | 0.65 | 0.78 | 0.85 | **0.84** |

Notes: CR = Composite Reliability; AVE = Average Variance Extracted; MSV = Maximum Shared squared variance.

**Table 9.** Items of the original tool that produced acceptable convergent and discriminant validity of the MEC App.

| Item | MEC App | Factor Loadings |
| --- | --- | --- |
| **Acceptability** |  |  |
| 1 | The MEC app is appealing to me | 0.60 |
| 2 | I like the MEC app | 0.81 |
| 3 | I welcome the MEC app | 0.84 |
| **Appropriateness** |  |  |
| 1 | The MEC app is suitable | 0.90 |
| 2 | The MEC app is applicable | 0.80 |
| 3 | The MEC app is a good match | 0.77 |
| **Feasibility** |  |  |
| 1 | The MEC app is implementable | 0.86 |
| 2 | The MEC app is possible | 0.87 |
| 3 | The MEC app is doable | 0.78 |

## Discussion

This study sought to assess the psychometric properties of the AIM, FIM, IAM when used to evaluate a one-on-one counselling intervention and the MEC App for the delivery of family planning services among midwives in Ghana. Specifically, we assessed the internal consistency, factor structure, and predictive validity, as well as the convergent validity and discriminant validity of the AIM, IAM, and FIM evaluating both one-on-one counselling and the MEC App intervention.

The mixed-method design was central to strengthening the psychometric performance of the adapted measures. Qualitative information obtained from providers clarified context-specific interpretations of acceptability, appropriateness, and feasibility, which guided the development of additional culturally relevant items. These adaptations improved the conceptual clarity of the measures and likely contributed to the strong internal consistency and improved model fit observed in the quantitative analyses.

The results of the study demonstrate that the internal consistency of the culturally adapted items measuring the acceptability (12 items), appropriateness (seven items), and feasibility (eight items) of one-on-one family planning counselling exceeded the minimum threshold. This suggests potential relevance of the adapted measures for similar provider populations in other low resource settings(23,24). These reliability coefficients are comparable to those reported in the original validation study by Weiner and colleagues, where Cronbach alpha values ranged between approximately 0.85 and 0.91 across the three implementation outcome measures. The similarity in reliability estimates suggests that the constructs of acceptability, appropriateness, and feasibility remain internally coherent when applied in this new service delivery context. Minor differences in reliability and factor loadings may reflect contextual variations in language, provider workflow, and familiarity with digital decision support tools, particularly in settings where mobile health technologies are still being integrated into routine clinical practice.

This is particularly relevant within the implementation outcomes framework proposed by Proctor and colleagues, which emphasises acceptability, appropriateness, and feasibility as early indicators of implementation success. Studies in other locations have shown similar patterns where original items work well, but adapted items sometimes demonstrate weaker construct validity. This raises questions about how far implementation outcomes can be assumed to generalize across settings without careful adaptation and testing. In this study, cultural adaptation improved relevance but did not always strengthen psychometric performance, which underscores the need for careful balance between contextual fit and measurement stability.

Regarding the AIM, IAM, and FIM tools in their evaluation of use of the MEC App, the results showed that the internal consistency of the culturally adapted items measuring the acceptability (13 items), appropriateness (seven items), and feasibility (seven items) of midwives using the mobile application to deliver family planning services is high. As with one-on-one counselling, our results support the finding that midwives in other low resource settings would also find the use of the MEC App acceptable, appropriate, and feasible (25). This may be because of the guidelines provided by the World Health Organization that emphasize the importance of using technology, including mobile applications, to enhance the delivery of family planning services, particularly in resource-constrained settings (26).

Results of the assessment of the factor structure of acceptability, appropriateness, and feasibility of one-on-one counselling and the MEC App for family planning showed that the culturally adapted scales have acceptable factor loadings, indicating that the AIM, IAM, and FIM measures are unidimensional assessing their respective underlying constructs for both one-on-one counselling and the MEC App. The convergence of qualitative and quantitative findings indicates that the adapted items reflected dimensions of implementation outcomes that were meaningful to providers’ routine practice. The unidimensional structure of the scales supports their internal coherence and interpretability in this context (27).

Findings from the predictive validity analysis showed that there was a positive association between the AIM, IAM, and FIM measures when used to assess one-on-one counselling. This positive association suggests that midwives who found one-on-one counselling to be more acceptable, appropriate, and feasible were more likely to prefer working in a facility where one-on-one counselling intervention is used. This finding suggests that one-on-one counselling, as an intervention, possesses good predictive validity in terms of influencing midwives’ preferences regarding their work environment. This aligns with previous research indicating that the perceived acceptability, appropriateness, and feasibility of an intervention are important factors affecting its adoption and success (14,28).

Similarly, the MEC App exhibits good predictive validity in terms of influencing midwives’ perceptions regarding the potential impact on their work efficiency. These findings align with the idea that technology acceptance and perceived usability are vital factors influencing the successful integration of digital tools in various sectors, including healthcare (27,29).

Finally, in the assessment of convergent and discriminant validity of the acceptability, appropriateness, and feasibility of one-on-one family planning counselling and the MEC App among midwives, the results showed good convergent validity and clear discriminant validity. These are based on the AIM, IAM, and FIM measures having four, three, and three items for acceptability, appropriateness, and feasibility respectively for one-on-one counselling and three items each for acceptability, appropriateness, and feasibility of the MEC App in the original outcome measures by Weiner and colleagues (14). This indicates that the items used to assess acceptability, appropriateness, and feasibility are distinct and do not overlap with each other. This is essential for accurately evaluating the effectiveness of the one-on-one counselling intervention and the MEC App. In other words, the AIM, IAM, and FIM have the capacity to proficiently distinguish midwives’ acceptability, appropriateness, and feasibility of delivering one-on-one counselling and using the MEC App for family planning counselling (30,31). Furthermore, this implies that when determining the suitability, relevance, and practicality of one-on-one family planning counselling and the MEC App among midwives, we can be confident in both assessing distinct aspects and avoiding confusion between the AIM, IAM, and FIM (14). However, the expanded versions of the culturally adapted AIM, IAM, and FIM did not consistently meet criteria for convergent and discriminant validity, whereas the abridged versions retained robust construct validity.

Some adapted items did not demonstrate stronger psychometric performance than the original items, likely because introducing additional items can alter the internal structure of a measure. Some adapted items captured language and expressions that providers used in practice, but these additions may also have introduced variation that reduced the statistical strength of the original four item structure. In other cases, the original items were already well aligned with how providers understood the constructs, so additional items did not improve performance. These patterns show that adaptation improves relevance but does not guarantee stronger statistical results, which supports the need to balance contextual fit with measurement stability during adaptation work.

Linking the qualitative findings with quantitative testing demonstrates that culturally adapted measures can maintain psychometric strength while improving contextual relevance. This integrated approach enhances confidence that the adapted AIM, IAM, and FIM scales are both analytically robust and practically meaningful for assessing postpartum family planning interventions in Ghana and similar settings.

The findings also contribute to implementation science by providing empirical evidence on the cross-cultural performance of widely used implementation outcome measures in a low- and middle-income country health system. This evidence is particularly important for implementation research conducted in low- and middle-income countries, where validated measurement tools are often applied across diverse health system contexts without prior psychometric assessment. To our knowledge, this study represents one of the first psychometric evaluations of the AIM, IAM, and FIM measures among midwives delivering postpartum family planning services in Ghana. The results demonstrate that frontline providers in this context are able to meaningfully evaluate both counselling interventions and digital decision support tools using these constructs, suggesting a level of digital readiness among midwives that may support future mobile health innovations.

### Limitations of the study

The study was conducted among midwives and family planning providers in Ghana, so, the results may not be generalizable to a broader population of healthcare providers or other geographical regions. The sample’s unique characteristics may limit the external validity of the findings. Moreover, midwives and family planning providers in different cultural and healthcare contexts may have different perceptions and experiences. In addition, formal cognitive interviewing and quantitative content validity indexing were not conducted during the adaptation process, which may limit the extent to which semantic equivalence of individual items can be fully assessed. Also, the cross-cultural adaptation process might not be perfect and so may not truly reflect the constructs of acceptability, appropriateness, and feasibility as intended in the Ghanaian context. The study did not include women receiving counselling, so client experiences of the one-on-one counselling approach or the MEC App were not captured. These perspectives may differ from those of providers and may add complementary information in future research. Also, the study relied on self-report data to assess the acceptability, appropriateness, and feasibility of the interventions. This might not accurately reflect the actual behaviours or practices of the midwives and family planning providers and may be influenced by social desirability. In addition, the absence of an independent validation sample limits the ability to perform cross validation of the factor structure, and the psychometric findings should therefore be interpreted as preliminary evidence of measurement performance in this context. Finally, test-retest reliability was not assessed because the survey was administered at a single time point following the implementation period. Future studies could examine the temporal stability of the adapted measures using repeated assessments.

## Conclusion

This study demonstrates that the original AIM, IAM, and FIM measures (14)require contextual adaptation to effectively capture provider experiences with one-on-one family planning counselling and use of the MEC App in Ghana. The adapted scales contribute to ongoing efforts in implementation science to refine and validate measurement tools that are both theoretically sound and practically relevant. While grounded in a specific intervention, these adaptations offer a foundation for scale development in similar reproductive and maternal health contexts, particularly those involving midwives and frontline providers in low-resource settings. This study has identified outcome measures that demonstrate good reliability and validity in assessing the acceptability, appropriateness, and feasibility of employing one-on-one family planning counselling in this specific context. These measures can be derived from either the abridged version of the modified original tool, which comprises four, three, and three items for acceptability, appropriateness, and feasibility, respectively, or from the expanded version in the present investigation, which encompasses twelve, seven, and eight items for acceptability, appropriateness, and feasibility, respectively.

For the assessment of the MEC App, reliable and valid outcome measures encompass three items each for acceptability, appropriateness, and feasibility in the original tool, and thirteen, seven, and seven items for acceptability, appropriateness, and feasibility, respectively, in the expanded version developed in this study. These findings support the use of the adapted AIM, IAM, and FIM measures for evaluating implementation outcomes in postpartum family planning programmes involving midwives and frontline providers in Ghana. The results also suggest that widely used implementation outcome instruments can retain strong psychometric performance when applied in new cultural and service delivery contexts, provided that careful adaptation and validation are undertaken.

## Data Availability

All data produced in the present study are available upon reasonable request to the authors

## Declarations

### Ethics approval and consent to participate

Ethical clearance for this study was obtained from the Ghana Health Service Ethics Review Committee (Protocol ID GHS-ERC 001/05/21). Written informed consent to participate was obtained from all participants prior to participation.

### Consent for publication

Not Applicable

### Availability of data and materials

The data used for this study are available upon reasonable request from the lead author.

### Authors’ contributions

EM, CG, FNG, SS, AM, RD, and MEG contributed to the conception and design of the work. EM, CG, contributed to the acquisition of the data. FNG, CG conducted data analysis. EM, CG FNG, SS, AM, RD, MEG and JK, contributed to the interpretation of the data. SS, EM, CG, FNG contributed to drafting the manuscript. All authors read and approved the final manuscript.

## Acknowledgments

The authors would like to acknowledge the contributions of our research assistants on this project. Additionally, we are grateful to the family planning fellow and our clinical leads who reviewed the modified tools for the survey. We would also like to thank the midwives and nurses who took part in this study. Again, we are grateful to the management and staff of all the health facilities used in the study. Finally, we would like to express our appreciation to the World Health Organization (A65990) and the University of Pennsylvania (Penn Presbyterian Harrison Award & RRP-4492402666) for providing the funds for this study.

